# Predictors of High-grade Squamous Intraepithelial Lesion treatment failure

**DOI:** 10.1101/2023.11.22.23298918

**Authors:** S. Botting-Provost, A. Koushik, H. Trottier, F. Coutlée, MH Mayrand

## Abstract

**Objective:** To estimate the association between several risk factors and high-grade squamous intraepithelial lesions (HSIL) treatment failure in order to identify predictors.

**Methods:** The study population included 1,548 Canadian women treated for HSIL who participated in a randomized control trial. HSIL treatment failure was the presence of histologically confirmed HSIL or worse during the two-year follow-up period. This nested-case control study included all 101 cases of treatment failure and controls that were matched 1:1 on treatment center and date of failure. Conditional logistic regression models were used to estimate odds ratios (ORs) and 95% confidence intervals (CIs) between each potential predictor and HSIL treatment failure. Independent variables that were examined included age, parity, smoking status, number of sexual partners, condom use, method of contraception, margins, number of passes, diagnosis on conisation, genotype, and number of infecting types. Interactions between smoking and margins and genotype were evaluated.

**Results:** Having positive vs. negative margins (adjusted OR=4.05, 95% CI 1.57-10.48) and being positive for *Human Papillomavirus* (HPV)16 and/or HPV18 vs. any other type (adjusted OR=2.69, 95% CI 1.32-5.49) were predictors of HSIL treatment failure in multivariable models. ORs suggested that older age, more severe lesions, and single-type infections may be at a higher risk of treatment failure but were not statistically significant. The ORs for smoking status, number of sexual partners, condom use, contraception, parity, and number of passes were near the null value. We did not observe any evidence of interaction between smoking and genotype, nor between margins and genotype.

**Conclusion:** Only positive margins and HPV16/18 positivity were predictors for being diagnosed with HSIL or worse within two years of treatment. However, we do not recommend automatic retreatment of those with positive margins because over 90% of those with positive margins did not fail treatment. The predictive value of HPV16 and HPV18 for HSIL treatment failure suggests that high coverage vaccination programs should contribute to a significant reduction in residual/recurrent disease.

## Introduction

It is estimated that 1,350 Canadian women will be diagnosed with cervical cancer and 410 women will die of it in 2020^1^. Globally, cervical cancer ranks fourth in terms of incidence and mortality amongst cancers affecting women^2^. Screening is a highly effective method of cervical cancer prevention^3–7^. Indeed, cervical cancer has a long pre-invasive phase that can be identified on histopathology, and the treatment of precancerous lesions, or High-grade squamous intraepithelial lesions (HSIL), reduces the incidence of cervical cancer^8^. Loop Electrosurgical Excision Procedure (LEEP) is the preferred treatment for cervical precancers or microinvasive cancers as it makes it possible to excise a limited and predetermined amount of cervical tissue and has the best success/side effect profile of conservative treatments^9^.

Despite efforts to best treat women diagnosed with HSIL, 10-15% of those treated will have recurrent or residual disease, known as treatment failure^10^. Women who have been treated for HSIL have 4-5 times the risk of developing cervical cancer as women in the general population ^11^. Those who experience treatment failure need to be retreated^12^. However, women who have undergone repeated excision of the cervix are at an increased risk of negative obstetric outcomes including second trimester pregnancy loss, preterm birth, and their offspring are at increased risk of low birth weight, complications of prematurity, and even neonatal death^12–14^. A large Danish cohort found that 33% of women with two conisations prior to pregnancy would experience preterm delivery^14^. In addition, preterm premature rupture of the membranes (pPROM) occurred in 92% of spontaneous preterm births in women with two prior conisations. Given the dire consequences, it is important to identify risk factors for treatment failure, as some may be modifiable and may help devise strategies to decrease treatment failure. Several patient, behavioural, clinical and viral risk factors for treatment failure have been studied^15–44^. Except for positive treatment margins that are strongly associated with treatment failure, evidence remains inconsistent for most risk factors. This is due to small sample sizes and a focus on a small number of potential risk factors simultaneously. Since exposures that are potentially associated with treatment failure are often highly correlated, results from past studies using univariate models may have potentially been biased, preventing the identification of independent predictors. Thus, the objective of our study was to explore a large number of potential risk factors for HSIL treatment failure, in order to identify predictors of treatment failure.

## Methods

### Study population

We used data collected in the Colposcopy vs. HPV testing to identify persistent precancers post treatment (CoHIPP) study, a randomized controlled trial (ClinicalTrials.gov Identifier: NCT01051895). CoHIPP methods have been described in detail previously [ref pending]. Briefly, women were recruited at the time of excisional treatment for HSIL (98% by LEEP); 1,548 had HSIL confirmed on the treatment specimen and were randomized to an HPV based follow-up strategy versus usual care. Participants in both groups were seen 6 months, 12 months, and 24 months post treatment. All participants had exocervical and endocervical biopsies at 12 and 24 months.

For the present analysis, we performed a nested case-control study within the CoHIPP cohort. A total of 101 cases of treatment failure were identified within the study population. Cases included all participants who had histologically confirmed HSIL or worse at any point during the two-year follow-up. Controls were selected using incidence density sampling and matched 1:1 to cases by treatment center and by visit, which had to have occurred within 3 months of the case’s identification. Controls were participants of the CoHIPP cohort who had not experienced treatment failure and who had not been censored at the time of matching. Of the 101 matched controls, one became a case at a later visit and nine were matched to two different cases.

CoHIPP received approval from the Research Ethics Committee of the Centre Hospitalier de l’Université de Montréal (CHUM). Participants provided free and informed consent to participate in CoHIPP and consented to the use of their data and specimens for additional studies on HPV and cervical precancers.

### Data collection

Socio-demographic and behavioural data were collected using self-administered questionnaires at each visit. The randomisation visit questionnaire was used to measure baseline characteristics and exposures. Cervical specimens were also collected at each visit and banked for further biomolecular analysis. For this analysis, we used the randomisation visit questionnaire data and the specimens that were collected just prior to treatment for all biomolecular viral analyses.

### Variables of interest

Variables considered included age, parity, smoking status, number of sexual partners in the last year, condom use in the last year, method of contraception, positivity of margins, number of passes, diagnosis on LEEP, genotype and number of types. The age of each participant was calculated at the time of treatment and was analysed as a continuous variable. Parity for this study was low and was categorised as 0 (reference), 1, and 2 or more. We compared current smokers to current non-smokers (reference) at randomisation. Number of sexual partners in the last year included current partners and we compared having 2 or more partners to having 0 or 1 partner (reference). Condom use in the last year was analysed as a binary variable comparing ever vs. never (reference). The method of contraception was also binary, comparing hormonal contraception which included oral contraceptives, hormonal intrauterine device (IUD), injectable contraceptive, hormonal patch, and vaginal ring, or any non-hormonal/hormonal combination, to non-hormonal or no contraception (reference), which included copper IUD, diaphragm, condoms, cervical cap, and no birth control. Margins on treatment specimen were analysed as a dichotomous variable comparing those with at least one positive margin to those with negative margins (reference). For number of loop passes done for treatment, we compared 2 or more passes to 1 pass (reference). Diagnosis on LEEP could have been the less severe Cervical Intraepithelial Neoplasia (CIN) 2, the more severe CIN3-Carcinoma in situ (CIS)-Adenocarcinoma in situ (AIS), or HSIL, not specified. Those with CIN2 were used as the reference group. HPV genotypes were detected on the cervical scrape collected immediately prior to treatment using Linear Array® assay from Roche Diagnostics, which identifies 37 types of HPV DNA^45^. Because of the clinical significance of both HPV16 and HPV18, a binary variable was created to compare those with HPV16 and/or HPV18 to infection with any other types (reference). Finally, we compared infection with 2 or more types to 1 type (reference).

### Statistical analysis

Statistical analyses were conducted using SAS version 9.4 for Windows (SAS Institute, Cary Inc.). Age was described by median, range, and interquartile range. Other variables were described by the number and proportion of cases or controls in each category. Odds ratios (OR) and 95% confidence intervals (CI) were estimated using conditional logistic regression models. Distinct univariable and multivariable models were developed for each potential predictor. The confounders included in each multivariable model had to be associated with both the independent variable of interest and the outcome variable. Due to the limited literature on predictors of HSIL treatment failure, there was an absence of known causal associations with our outcome. We therefore could not rely on the use of Directed Acyclic Graphs (DAG) to identify confounders that should be included in our multivariable models. In fact, we suspected that any one of our independent variables may be strongly associated with treatment failure. Any variable that was associated with the predictor of interest could then be considered as a potential confounder and included in the multivariable model. We suspected that many of our potential predictors were correlated^24^ and neglecting to account for these correlations would have led to confounding bias. To determine the strength of these correlations, we estimated Spearman correlation coefficients between all of our independent variables. For each predictor, we included covariates for which the absolute value of the Spearman correlation coefficient was ≥0.2, as done previously^46^. Age was forced into all multivariable models, regardless of the strength of the correlation.

To avoid dropping of matched pairs from the analysis in cases where confounders contained missing data, we imputed the most frequent value among cohort members to replace missing data within the confounders of each model. However, in the main analyses of each variable of interest, the variable was left as-is in order to preserve the observed distribution. To ensure that this method did not overly bias the multivariable ORs, we conducted a sensitivity analysis comparing multivariable ORs and 95% CIs with and without the use of imputation in the confounder variables. Matched pairs with missing values were dropped from the analysis without imputation.

## Results

**Table 5.1** summarises the baseline characteristics of the study population. The median age was 35.28 for cases and 30.15 for controls. Over 50% of the population was nulliparous, specifically 48.51% of cases and 55.45% of controls. A slightly higher proportion of cases were current smokers than controls. The population was largely monogamous with over 70% of both cases and controls reporting 0 or 1 sexual partner in the last year. Most participants never used condoms, likely because the group was largely monogamous. Distribution of condom use in the last year was almost the same for cases and controls, with 44.6% of cases and 47.5% of controls never using condoms. Fewer cases than controls used hormonal contraception. Far more cases had at least one positive margin (55.5%) compared to controls (33.7%). The distribution of number of passes was identical in cases and controls. More cases than controls had a more severe diagnosis of CIN3-CIS-AIS on LEEP, and HSIL type was not specified for 40.6% of cases and 34.7% of controls. Only 11.9% (24/202) of the study population had received an anti-HPV vaccine. Furthermore, given the age of the population and the recruitment period, most participants who were vaccinated received their first dose several years after initiation of sexual activity, when effectiveness is reduced ^47,48^. The variable was thus not analysed any further. As expected, more cases than controls had HPV16 and/or HPV18 at baseline. More controls than cases were infected with multiple types of HPV. All variables had less than 5% missing data except for condom use, method of contraception and margins, which had 11.9% (14 cases; 10 controls), 11.4% (12 cases; 11 controls) and 20.8% (23 cases; 19 controls) missing values respectively.

**Table 5.1.**
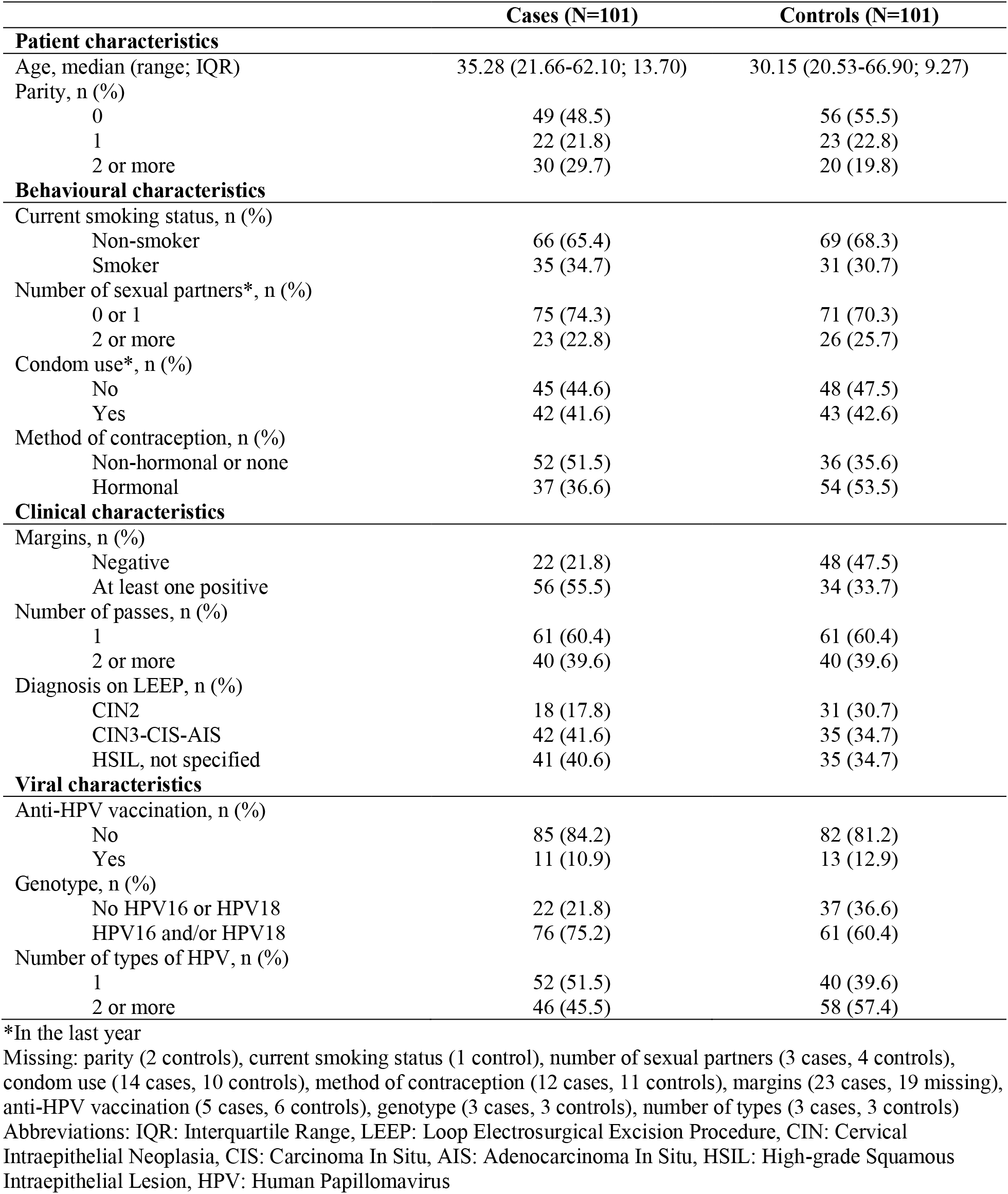
Baseline characteristics of study population.

|  | Cases (N=101) | Controls (N=101) |
| --- | --- | --- |
| **Patient characteristics** |  |  |
| Age, median (range; IQR) | 35.28 (21.66-62.10; 13.70) | 30.15 (20.53-66.90; 9.27) |
| Parity, n (%) |  |  |
| 0 | 49 (48.5) | 56 (55.5) |
| 1 | 22 (21.8) | 23 (22.8) |
| 2 or more | 30 (29.7) | 20 (19.8) |
| **Behavioural characteristics** |  |  |
| Current smoking status, n (%) |  |  |
| Non-smoker | 66 (65.4) | 69 (68.3) |
| Smoker | 35 (34.7) | 31 (30.7) |
| Number of sexual partners*, n (%) |  |  |
| 0 or 1 | 75 (74.3) | 71 (70.3) |
| 2 or more | 23 (22.8) | 26 (25.7) |
| Condom use*, n (%) |  |  |
| No | 45 (44.6) | 48 (47.5) |
| Yes | 42 (41.6) | 43 (42.6) |
| Method of contraception, n (%) |  |  |
| Non-hormonal or none | 52 (51.5) | 36 (35.6) |
| Hormonal | 37 (36.6) | 54 (53.5) |
| **Clinical characteristics** |  |  |
| Margins, n (%) |  |  |
| Negative | 22 (21.8) | 48 (47.5) |
| At least one positive | 56 (55.5) | 34 (33.7) |
| Number of passes, n (%) |  |  |
| 1 | 61 (60.4) | 61 (60.4) |
| 2 or more | 40 (39.6) | 40 (39.6) |
| Diagnosis on LEEP, n (%) |  |  |
| CIN2 | 18 (17.8) | 31 (30.7) |
| CIN3-CIS-AIS | 42 (41.6) | 35 (34.7) |
| HSIL, not specified | 41 (40.6) | 35 (34.7) |
| **Viral characteristics** |  |  |
| Anti-HPV vaccination, n (%) |  |  |
| No | 85 (84.2) | 82 (81.2) |
| Yes | 11 (10.9) | 13 (12.9) |
| Genotype, n (%) |  |  |
| No HPV16 or HPV18 | 22 (21.8) | 37 (36.6) |
| HPV16 and/or HPV18 | 76 (75.2) | 61 (60.4) |
| Number of types of HPV, n (%) |  |  |
| 1 | 52 (51.5) | 40 (39.6) |
| 2 or more | 46 (45.5) | 58 (57.4) |

\*In the last year
Missing: parity (2 controls), current smoking status (1 control), number of sexual partners (3 cases, 4 controls), condom use (14 cases, 10 controls), method of contraception (12 cases, 11 controls), margins (23 cases, 19 missing), anti-HPV vaccination (5 cases, 6 controls), genotype (3 cases, 3 controls), number of types (3 cases, 3 controls)
Abbreviations: IQR: Interquartile Range, LEEP: Loop Electrosurgical Excision Procedure, CIN: Cervical Intraepithelial Neoplasia, CIS: Carcinoma In Situ, AIS: Adenocarcinoma In Situ, HSIL: High-grade Squamous Intraepithelial Lesion, HPV: Human Papillomavirus

**Table 5.2** shows the frequency of each HPV genotype in the LEEP specimens. High-risk types of HPV were detected more frequently than low-risk types among both cases and controls. HPV16 was the most frequent type in both cases and controls, followed by HPV31 and HPV52. The next most frequent types in cases were HPV53, HPV18 and HPV89, while in controls they were HPV33 and HPV62. 73 cases had HPV16 compared to 59 controls. There were only 11 participants infected with HPV18 (6 cases, 5 controls).

**Table 5.2.** Frequency of HPV genotypes in LEEP specimens.

| **HPV Genotype** | **Cases (N=101)** | **Controls (N=101)** |
| --- | --- | --- |
| **High-risk types** | **137\*** | **144** |
| 16 | 73 | 59 |
| 18 | 6 | 5 |
| 31 | 13 | 17 |
| 33 | 4 | 12 |
| 35 | 3 | 6 |
| 39 | 3 | 4 |
| 45 | 2 | 6 |
| 51 | 5 | 7 |
| 52 | 13 | 16 |
| 56 | 5 | 2 |
| 58 | 5 | 3 |
| 59 | 5 | 7 |
| **Low-risk types** | **55** | **57** |
| 6 | 3 | 1 |
| 34 | 0 | 1 |
| 40 | 1 | 1 |
| 42 | 2 | 4 |
| 44 | 1 | 3 |
| 53 | 7 | 3 |
| 54 | 6 | 6 |
| 61 | 4 | 1 |
| 62 | 6 | 12 |
| 66 | 2 | 1 |
| 67 | 2 | 4 |
| 68 | 1 | 0 |
| 69 | 0 | 1 |
| 73 | 1 | 3 |
| 81 | 2 | 2 |
| 82 | 3 | 3 |
| 83 | 3 | 2 |
| 84 | 5 | 4 |
| 89 | 6 | 5 |

\*Total frequency of genotypes is greater than N due to the presence of co-infection, or infection with multiple types of HPV in one participant Missing: 3 cases, 3 controls

In our primary analysis (**Table 5.3**), we found that age, parity, being a smoker, number of sexual partners, use of condoms or hormonal contraception were not associated with the risk of treatment failure. As expected, having positive margins was a strong predictor of treatment failure (OR 4.05, 95% CI 1.57-10.48); however, the number of passes was not. Compared to a CIN2 diagnosis on LEEP, the ORs for treatment failure was 1.63 (95% CI 0.76-3.49) for CIN3-CIS-AIS and 2.12 (95% CI 0.81-5.55) for unspecified HSIL, but neither were statistically significant. Positivity for HPV16 or 18 vs. other HPV types at the time of LEEP was associated with a higher risk of treatment failure. On the other hand, the OR for having infection with multiple HPV types vs. a single type was 0.43 with 95% CI 0.17-1.09. When analyses were restricted to pairs with complete data (i.e. covariate values not imputed), the results presented in Table 3 did not greatly change (**Supplementary table 5.1**). Imputation of the most frequent value decreased the variability of our data. Confidence intervals therefore tended to be slightly narrower in analysis with imputation compared to without.

**Table 5.3.** Univariable and multivariable analysis of potential predictors for HSIL treatment failure.

| Potential predictor | Crude OR<br>(95% CI) | Adjusted OR<br>(95% CI) | Covariates in adjusted model |
| --- | --- | --- | --- |
| Age, per 5-unit increase | 1.28 (1.10-1.47) | 1.28 (0.90-1.76) | Parity, number of sexual partners, condom use, method of contraception, number of types |
| Parity |  |  |  |
| 0 | 1 | 1 | Age, number of sexual partners, condom use |
| 1 | 1.07 (0.53-2.18) | 0.49 (0.19-1.34) |  |
| 2 or more | 1.63 (0.84-3.17) | 0.87 (0.31-2.45) |  |
| Current smoking status |  |  |  |
| Non-smoker | 1 | 1 | Age, number of passes |
| Smoker | 1.67 (0.62-2.19) | 1.02 (0.51-2.05) |  |
| Number of sexual partners* |  |  |  |
| 0 or 1 | 1 | 1 | Age, parity, condom use, number of types, margins |
| 2 or more | 0.82 (0.40-1.67) | 1.16 (0.41-3.29) |  |
| Condom use* |  |  |  |
| No | 1 | 1 | Age, parity, number of sexual partners |
| Yes | 1.06 (0.55-2.01) | 1.32 (0.56-3.09) |  |
| Method of contraception |  |  |  |
| Non-hormonal or none | 1 | 1 | Age |
| Hormonal | 0.48 (0.24-0.96) | 0.74 (0.34-1.61) |  |
| Margins |  |  |  |
| Negative | 1 | 1 | Age, number of sexual partners, number of passes |
| At least one positive | 3.75 (1.72-8.18) | 4.05 (1.57-10.48) |  |
| Number of passes |  |  |  |
| 1 | 1 | 1 | Age, current smoking status, margins |
| 2 or more | 1.00 (0.53-1.89) | 0.94 (0.45-1.97) |  |
| Diagnosis on LEEP |  |  |  |
| CIN2 | 1 | 1 | Age |
| CIN3-CIS-AIS | 1.96 (0.94-4.07) | 1.63 (0.76-3.49) |  |
| HSIL, not specified | 2.43 (0.95-6.23) | 2.12 (0.81-5.55) |  |
| Genotype |  |  |  |
| No HPV16 or HPV18 | 1 | 1 | Age, number of types |
| HPV16 and/or HPV18 | 2.21 (1.18-4.16) | 3.03 (1.44-6.41) |  |
| Number of types of HPV |  |  |  |
| 1 | 1 | 1 | Age, number of sexual partners, genotype |
| 2 or more | 0.57 (0.29-1.12) | 0.43 (0.17-1.09) |  |

\*In the last year
OR: Odds Ratio, CI: Confidence Interval, LEEP: Loop Electrosurgical Excision Procedure, CIN: Cervical Intraepithelial Neoplasia, CIS: Carcinoma In Situ, AIS: Adenocarcinoma In Situ, HSIL: High-grade Squamous Intraepithelial Lesion, HPV: Human Papillomavirus

Finally, we explored effect modification between key variables. Smoking may reduce one’s immunity to infection with HPV16, decreasing the number of circulating antibodies ^49^. We hypothesised that being a current smoker could modify the oncogenic effect of HPV16/HPV18 and increase the risk of HSIL treatment failure. However, no interaction was found between genotype and smoking status (**Supplementary table 5.2**). Similarly, we hypothesised that the odds of treatment failure amongst those with HPV16 and/or HPV18 could be increased by the presence of positive margins. Again, no interaction was identified.

## Discussion

In our study of the potential risk factors for HSIL treatment failure, we found that positive margins and having HPV16 and/or HPV18 were significant predictors of HSIL treatment failure. The association between positive clinical margins and treatment failure was expected, and consistent with previous findings^16,17,20,22,28–30,33,36,50–53^. Overall, in CoHIPP, 10% of women with positive margins were diagnosed with HSIL within 2 years of treatment. Most likely these diagnoses represent persistent disease, as a positive margin indicates that some disease was left *in situ*. However, most women with positive margins will not have treatment failure. It is indeed possible for the disease to come to the edge of the cauterized region of the excised tissue, thus leading to a positive margin diagnosis, but for the whole lesion to be removed. It is also possible that the immune reaction secondary to the treatment injury could lead to the clearance of the HPV infection and small residual disease.

For larger lesions, surgeons can opt to use a smaller loop and perform multiple passes in order to avoid an excessively deep excision that would result from using a larger loop ^54,55^. This type of procedure can make staging impossible if a small invasive cancer is found on multiple pieces of the LEEP specimen, and thus would have a negative impact on the patient’s treatment plan^56^. However, it is reassuring that treatment with multiple passes was not associated with an increased risk of treatment failure.

In addition, those with HPV16 and/or HPV18 identified on their LEEP specimen had a significantly higher odds of treatment failure than those with all other HPV types identified by Linear Array. Our findings add to evidence previously found by Wu et al. who showed that single-type infections with HPV16, 18, 33 and 45 were associated with an increased risk of biopsy proven residual/recurrent disease^41^. In contrast to our study, theirs was limited to patients with negative margins on treatment. Nonetheless, HPV genotype was not associated with the positivity of margins in our study population (Spearman correlation = -0.13).

In a previous study, infection with multiple HPV types vs. single type infection was significantly associated with a greater risk of recurrent/residual disease^41^. In contrast, our results suggested a protective effect of being infected with multiple types, though the OR was not statistically significant. We would have expected that adjusting for genotype would attenuate this effect, since infections with HPV16 and/or HPV18 were more likely to be single-type infections. However, the OR for infection with multiple types compared to one type was 0.43 with 95% CI 0.17-1.09, even after adjustment. It is possible that this result is unique to our study population. Overall, our results suggested that the type present was a more significant predictor of treatment failure than the number of types.

Advanced age, especially being 50 years of age and older, is generally considered a risk factor for treatment failure^24–27,30,32^. Our findings suggested that the odds of treatment failure may indeed increase with age. For example, in univariable analysis, participants 40 years of age and older had 2.62 (95% CI 1.31-5.25) times the odds of treatment failure compared to participants under the age of 30. However, the adjusted odds measured per five-year increase in age were not significant in our study population, possibly owing to a small number of women of older ages. We also explored using 40 and 50 years and older as thresholds but did not find any significant association with the risk of treatment failure (**Supplementary Table 5.3**).

Although parity and use of hormonal contraception have been associated with a higher risk of cervical cancer in HPV positive women^57^ we, as others^27,28,32^ have not found these characteristics to be associated with HSIL treatment failure. We should note that our study population was generally of low parity, with only 2% of women reporting more than 3 deliveries. As such, we could not investigate the potential impact of higher parity.

In our study, current smoking status was not associated with treatment failure. This finding is in agreement with a small study that found that being a smoker and number of cigarettes smoked were not significantly associated with relapse of CIN^43^. They found, however, that being a smoker in conjunction with HPV positivity after treatment increased the risk of relapse with a higher grade of disease (CIN3 and microinvasive cancer). It is important to note that HPV positivity post-treatment may act as a surrogate measurement for the outcome of interest, biasing their results. In addition, they did not adjust for confounders. In contrast, a more robust prospective study of 77 cases of treatment failure and 154 controls that investigated the association between smoking and treatment failure found that, not only does being a current smoker increase the odds of treatment failure (OR= 3.17, 95% CI 1.68-5.91), but there is an observable dose-response relationship^15^. They estimated that for every additional 10 cigarettes smoked per day (from 0-30 cigarettes), the odds of treatment failure increased by a factor of 2.58 (95% CI 1.70-3.91). Their estimates were adjusted for HPV infection post-treatment, but not for potential sociodemographic confounders. In our study, only 33% of participants were current smokers, a smaller proportion than the other study populations (52% and 54%). In addition, with the data at our disposal, we were not able to quantify smoking in terms of cigarettes or packs per day and therefore did not measure the effect of dose. On the other hand, we adjusted for behavioural confounders that were strongly correlated with smoking within our population (number of sex partners and method of contraception) but were not accounted for in the other studies that investigated smoking and treatment failure. Associations observed between smoking status and treatment failure in previous studies may simply have been the result of confounding bias.

Diagnosis on LEEP, or increased severity of the lesion did not show a positive association with treatment failure. However, the OR for those with CIN3-CIS/AIS does suggest the possibility of an increased odds of treatment failure compared to those with CIN2. Judging by the even greater OR for the category HSIL not specified, we suppose that this group was primarily composed of participants with CIN3-CIS/AIS. Our analysis of this potential risk factor was limited by the proportion of subjects whose grade of lesion was not specified and this risk factor requires further investigation.

Despite being larger than most prior studies to have investigated predictors of treatment failure, this study was still limited by sample size, with only 101 cases of treatment failure identified in the CoHIPP cohort. This limits the statistical power of our analyses. The use of matched controls and conditional logistic regression should have produced estimates very similar to those that would have been obtained on the entire cohort. There was little missing data overall, however relative risks may have been biased for condom use, method of contraception and margins, which had 11.88%, 11.39% and 20.79% missing data respectively. In addition, our study was limited by the fact that the baseline questionnaire occurred 6 months after treatment. In fact, for condom use and number of sex partners, “in the last year” included 6 months prior to treatment and not a full year. This could have also affected measurement of current smoking status, since participants may have modified their behaviour since treatment. We also included a larger number of potential predictors than prior studies, which allowed us to control for potential confounding bias that had not been accounted for in other studies.

In conclusion, in this large cohort of unselected women who underwent treatment for HSIL, only having positive margins at treatment and being HPV16/18 positive were significantly associated with being diagnosed again with HSIL within two years. However, given that 90% of women with positive margins in CoHIPP did not experience treatment failure, and because of the risk to future pregnancies with repeated treatments^12–14^ we do not recommend automatic retreatment of women with positive treatment margins. Rather, our results emphasise the importance of mechanisms to minimize losses to follow-up in this group. Finally, given the singular role of HPV16/18 in HSIL treatment failure, the implementation of high coverage HPV vaccine programs should lead to a significant decrease in re-treatments, and limit the associated adverse obstetric impacts.

## Data Availability

All data in the present study is confidential as participants did not consent to data sharing.

---

**Supplementary Table 5.1.** Multivariable analysis without imputation*.

| Potential predictor | Cases<br>(N=101)<br>n (%) | Controls<br>(N=101)<br>n (%) | Adjusted OR<br>without imputation<br>(95% CI) | Covariates in adjusted<br>model |
| --- | --- | --- | --- | --- |
| Age, per 5-unit increase | 84 (83.2) | 85 (84.2) | 1.06 (0.97-1.15) | Parity, number of sexual<br>partners, condom use,<br>method of contraception,<br>number of types |
| Parity |  |  |  |  |
| 0 | 44 (43.6) | 51 (50.5) | 1 | Age, number of sexual<br>partners, condom use |
| 1 | 17 (16.8) | 21 (20.8) | 0.63 (0.19-2.04) |  |
| 2 or more | 24 (23.8) | 14 (13.9) | 1.11 (0.30-4.17) |  |
| Current smoking status |  |  |  |  |
| Non-smoker | 66 (65.3) | 69 (68.3) | 1 | Age, number of passes |
| Smoker | 35 (34.7) | 31 (30.7) | 1.02 (0.51-2.05) |  |
| Number of sexual partners<sup>+</sup> |  |  |  |  |
| 0 or 1 | 63 (62.4) | 62 (61.4) | 1 | Age, parity, condom use,<br>number of types, margins |
| 2 or more | 22 (21.8) | 24 (23.8) | 0.85 (0.62-4.49) |  |
| Condom use<sup>+</sup> |  |  |  |  |
| No | 44 (43.6) | 44 (43.6) | 1 | Age, parity, number of<br>sexual partners |
| Yes | 41 (40.6) | 42 (41.6) | 1.20 (0.50-2.88) |  |
| Method of contraception |  |  |  |  |
| Non-hormonal or none | 52 (51.5) | 36 (35.6) | 1 | Age |
| Hormonal | 37 (36.6) | 54 (53.5) | 0.74 (0.34-1.61) |  |
| Margins |  |  |  |  |
| Negative | 22 (21.8) | 42 (41.6) | 1 | Age, number of sexual<br>partners, number of passes |
| At least one positive | 48 (47.5) | 28 (27.7) | 3.71 (1.44-9.55) |  |
| Number of passes |  |  |  |  |
| 1 | 19 (18.8) | 16 (15.8) | 1 | Age, current smoking status,<br>margins |
| 2 or more | 12 (11.9) | 15 (14.9) | 0.96 (0.47-2.03) |  |
| Diagnosis on LEEP |  |  |  |  |
| CIN2 | 18 (17.8) | 31 (30.7) | 1 | Age |
| CIN3-CIS-AIS | 42 (41.6) | 35 (34.7) | 1.63 (0.76-3.49) |  |
| HSIL, not specified | 41 (40.6) | 35 (34.7) | 2.12 (0.81-5.55) |  |
| Genotype |  |  |  |  |
| No HPV16 or HPV18 | 22 (21.8) | 37 (36.6) | 1 | Age, number of types |
| HPV16 and/or HPV18 | 76 (75.2) | 61 (60.4) | 2.69 (1.31-5.49) |  |
| Number of types of HPV |  |  |  |  |
| 1 | 51 (50.4) | 40 (39.6) | 1 | Age, number of sexual<br>partners, genotype |
| 2 or more | 44 (43.6) | 56 (55.4) | 0.53 (0.21-1.37) |  |

\*Matched pairs with missing data were dropped from analysis
<sup>+</sup>In the last year
OR: Odds Ratio, CI: Confidence Interval, LEEP: Loop Electrosurgical Excision Procedure, CIN: Cervical Intraepithelial Neoplasia, CIS: Carcinoma In Situ, AIS: Adenocarcinoma In Situ, HSIL: High-grade Squamous Intraepithelial Lesion, HPV: Human Papillomavirus

**Supplementary Table 5.2.**
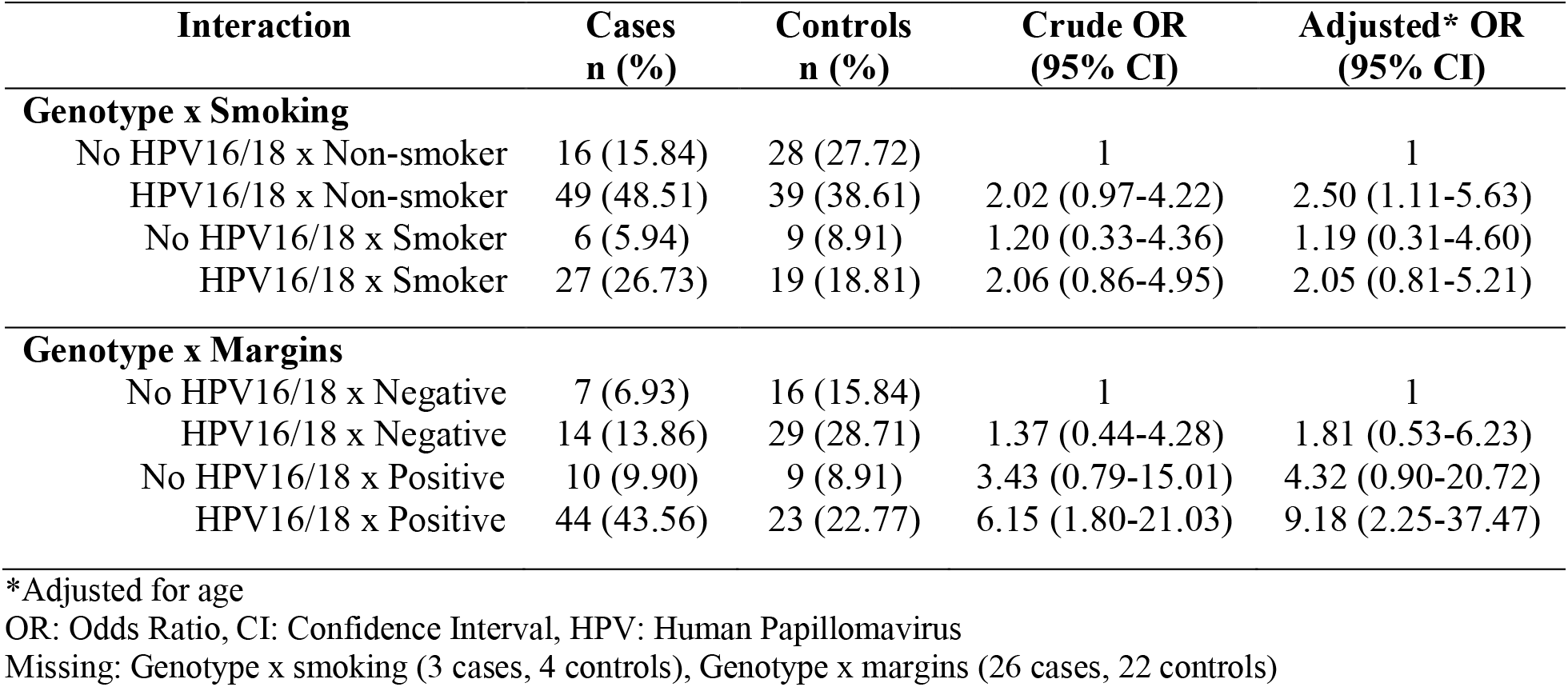
Effect of the interactions between smoking and margins with genotype.

**Supplementary Table 5.3.**
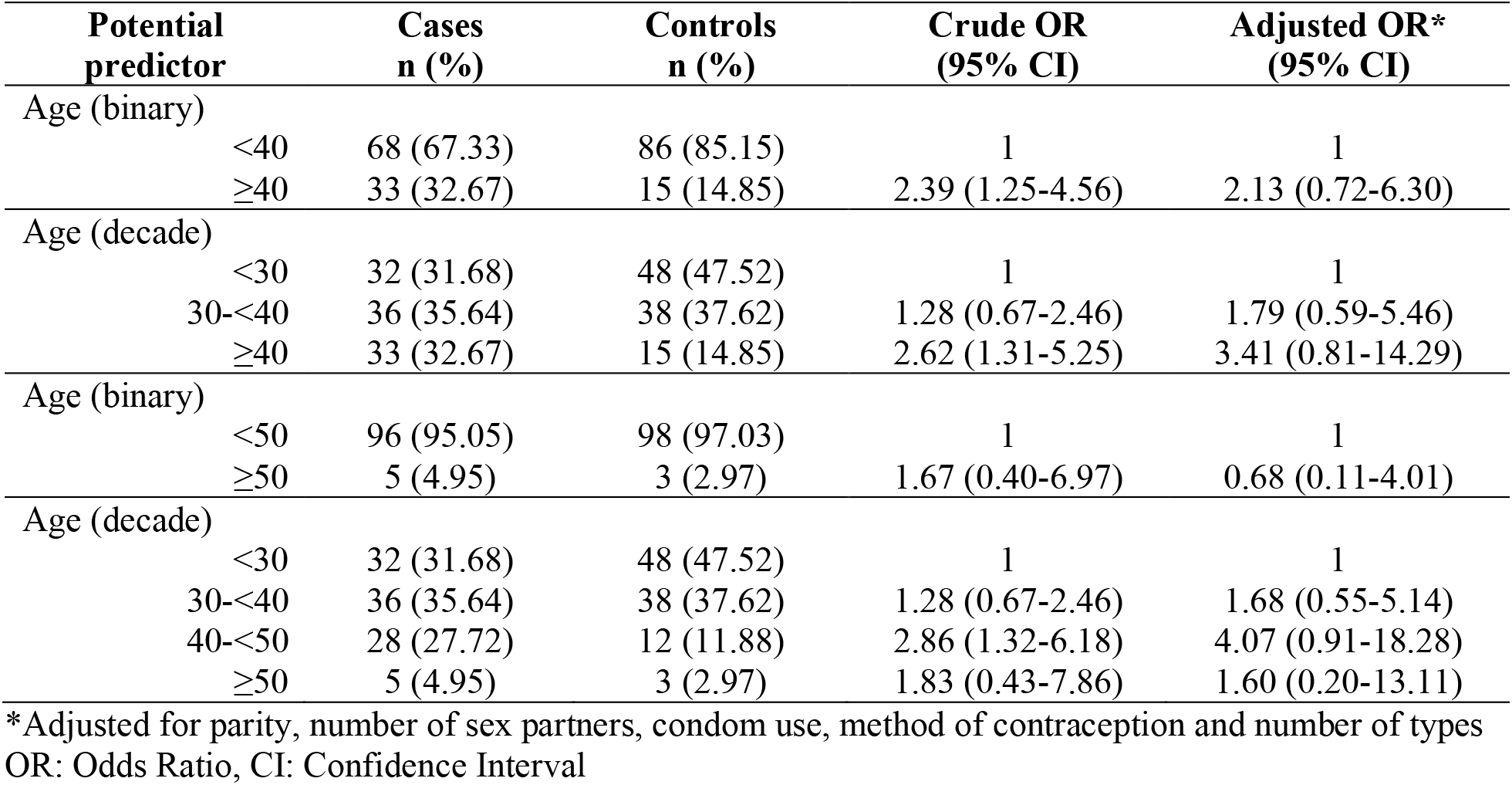
Age analyzed as a categorical variable.

## Notes

### Competing Interest Statement

The authors have declared no competing interest.

### Clinical Protocols

https://clinicaltrials.gov/study/NCT01051895

### Funding Statement

This study was funded by the Canadian Institutes for Health Research

### Author Declarations

Research Ethics Committee of the Centre Hospitalier de lUniversité de Montréal gave ethical approval for this work

